# Persistently uneven deep brain stimulation management in Japan: a nationwide claims analysis, 2014–2024

**DOI:** 10.64898/2026.09.14.26362780

**Authors:** Jun Tanimura, Takao Hashimoto

**Affiliations:** Department of Neurology, Aizawa Hospital, 2-5-1 Honjo, Matsumoto, Nagano 390-0814, Japan; Emory National Biomedical Research Center, Emory University, 954 Gatewood Road NE, Atlanta, GA 30329, USA

**Keywords:** deep brain stimulation, DBS programming, regional variation, claims data

## Abstract

**Background:** The long-term benefit of deep brain stimulation (DBS) for neurological disorders depends on sustained post-implantation management, yet its geographic distribution remains poorly characterised. We aimed to describe regional variation, persistence, and longitudinal change in DBS management activity across Japan.

**Methods:** We used claims from the National Database of Health Insurance Claims (NDB) Open Data during 2014–2024. Annual billings for DBS management were expressed per million population. Annual between-prefecture unevenness was summarised by the Gini coefficient of the prefecture DBS management billing rate. We evaluated longitudinal trajectories and trends in DBS management billing rates relative to the national rate, and related baseline billing rates to subsequent trends.

**Findings:** From 2014 to 2024, national DBS management billings increased from 18,659 to 27,233, totalling 258,408. Between-prefecture unevenness remained high throughout (Gini coefficient, 0.54 to 0.40), and its narrowing decelerated over the study period. In 2024, the prefecture at the 90th percentile still billed at 9.1 times the rate of the prefecture at the 10th. Of the 23 low-baseline prefectures, nine showed a subsequent decline in relative billing rate; nonetheless, lower baseline DBS management billing rates were associated with a more positive relative rate of change overall.

**Interpretation:** Ongoing post-implantation DBS management activity was markedly uneven and persistent across Japan. The observations suggest that universal health coverage and national-level growth do not by themselves ensure equitable long-term DBS treatment. This nationwide, longitudinal description could inform policies aimed at reducing geographic barriers to long-term specialist care across healthcare systems

**Funding:** None.

## Introduction

Deep brain stimulation (DBS) is a standard treatment for advanced Parkinson’s disease, essential tremor, and dystonia, and an option for selected patients with drug-resistant focal epilepsy^1,2^, with more than 230,000 devices estimated to have been implanted worldwide by 2021^3^. However, evidence suggests that DBS remains underused among patients who could potentially benefit^4,5^. Effective delivery of DBS treatment requires not only access to implantation surgery but also systems for longitudinal post-implantation management, in which neurologists, neurosurgeons, and specialised nurses collaborate to optimise stimulation settings through repeated adjustment^6,7^. Provision of DBS therefore cannot be understood from the implantation alone; it must also consider on-going management and follow-up.

Previous research on DBS has documented disparities in the receipt of implantation surgery by race, gender, income, insurance status, and geography^5,8–10^. These studies, however, have rarely been extended beyond surgery to routine, long-term post-implantation management^11^. In addition, because most rely on inpatient databases or provider cohorts, they characterise which individuals receive DBS in the cohort rather than how health systems distribute DBS services. Regional analysis, especially when it corresponds to administrative units, can instead directly inform healthcare resource optimisation within a nation^12^. In Japan, the prefecture is such a relevant regional unit^13^, providing an appropriate basis for comparing the nationwide delivery status of health services.

Japan faces the same knowledge gap in the equitable delivery of DBS management, and its health-system setting potentially offers lessons relevant across broader healthcare systems. Its universal health coverage and nationally uniform fee schedule reduce variation attributable to insurance entitlement and reimbursement arrangements^14^; thus observed prefectural differences more plausibly reflect how services are delivered. Because coverage is universal, national claims data further permit comprehensive comparison of billed DBS management activity. This comparison has gained new relevance since reimbursement for remote DBS management was introduced in 2026^15^, creating a need for explicit pre-policy benchmarks.

We therefore used all 11 years of the published National Database of Health Insurance Claims (NDB) Open Data (2014–2024) to characterise regional variation, persistence, and longitudinal dynamics in ongoing DBS management activity across Japan. To our knowledge, this is the first nation-wide longitudinal assessment of the geographic distribution of routine post-implantation DBS management in any nation. By characterising this variation and its persistence over time, this study would inform policies for reducing geographic barriers to long-term specialist care.

## Methods

### Study design and data source

We conducted a nationwide longitudinal ecological study using the NDB Open Data released by the Ministry of Health, Labour and Welfare^16^. We included all 47 prefectures and all 11 annual releases available at the data cut-off (July 20, 2026), covering fiscal year (FY) 2014 through FY2024. Because all available prefectures and years were included for descriptive purposes, no sample-size calculation was performed. Each release reports annual counts for each billing code over one fiscal year (April–March). To preserve comparability with the first nine releases, we excluded claims billed solely under public funding. The billing-code system and NDB Open Data source are described in the Supplementary Code Guide.

### Indicator for DBS management activity

The primary measure was the annual number of C110-2 billings, a claims measure for ongoing post-implantation DBS management. Ongoing DBS management indicates the repeated adjustment of stimulation settings and associated clinical follow-up, typically conducted in an outpatient setting but occasionally requiring inpatient care in more complex cases. The majority of patients billed under this code have movement disorders (see Supplementary Code Guide). The NDB Open Data applies small-cell suppression; prefecture-year counts below 10 are suppressed and published as blank cells. When exactly one cell (prefecture) in a given year is suppressed, the entire set of prefecture values for that year is suppressed and only the total is shown. This rule accounts for the FY2016 all-prefecture suppression of C110-2. We additionally used counts of patients with at least one C110-2 billing (the ID5-linked count), available only for FY2022–FY2024, to validate the correlation between C110-2 billings and patient counts (see Supplementary Code Guide).

### Statistical analysis

Annual DBS management billing rates were calculated for each prefecture by dividing billings by the corresponding population (October 1 estimate, Statistics Bureau of Japan) and were expressed per million population. Between-prefecture unevenness in the billing rate was summarised by the Gini coefficient ^17^ and by the ratio of its 90th to its 10th percentile. Each prefecture was given equal weight in the primary Gini analysis, reflecting our focus on prefectures as units of health-service planning. For comparison, we computed the equivalent Gini coefficient for prefecture-level neurologist, neurosurgeon, and total physician density (2018/2020/2022/2024 mean; e-Stat Physician, Dentist and Pharmacist Statistics). Population and physician workforce data sources are detailed in the Supplementary Methods. To characterise longitudinal change relative to the national trend, we divided each prefecture’s DBS management billing rate by the national billing rate in the same fiscal year; we refer to this ratio as the relative DBS management billing rate. Expressed as a percentage, it is equivalent to the standard-ised claim ratio computed without stratification, the measure conventionally used in Japanese claims data^18,19^. For each prefecture, we calculated the longitudinal trend in the relative billing rate as the Theil–Sen slope against fiscal year (the median of the slopes from every pair of available yearly values), expressed as percentage points of the national rate per year, with a rank-based 95% confidence interval.

We related each prefecture’s mean FY2014–FY2015 billing rate (the baseline billing rate) to its FY2017–FY2024 trend in relative billing rate (the subsequent trend). We separated the baseline and trend windows to avoid mathematical coupling and to bypass the FY2016 suppression gap. We categorised prefectures into three groups: no clear catch-up trend, for those with a baseline below the national mean and a negative subsequent trend; catch-up trend, for those with a baseline below the national mean and a non-negative subsequent trend; and high baseline, for those at or above the national mean. Spearman correlation (*ρ*) included prefectures with both baseline years and at least six subsequent observations. A 95% confidence interval for *ρ* used 20,000 prefecture bootstrap resamples. P value was calculated with a two-sided permutation test (100,000 permutations of the prefecture pairing between baseline and subsequent trend).

In an exploratory analysis, we related recent DBS management billing rate and its subsequent relative trend to five prefecture-level indicators: DBS implantation surgery billing (K181, see Supplementary Code Guide), neurologist, neurosurgeon, and total physician density (defined above), and per-capita prefectural income (FY2018–FY2022 mean, the most recent five years available; Cabinet Office^20^) as an indicator of regional economic strength. Effect sizes were assessed by Cliff’s *δ* for the binary DBS surgery billing indicator and Spearman’s *ρ* for the four continuous indicators, with 95% CI from 20,000 prefecture bootstrap resamples. Nominal P values were calculated with a two-sided permutation test (100,000 permutations), without adjustment for multiple comparisons.

Sensitivity and robustness analyses are detailed in the Supplementary Methods. Analyses were performed using Python 3.12. All maps used Natural Earth prefecture boundaries.

### Ethical statement

All data were publicly available summary counts at the prefecture or national level; the study therefore did not require ethics-committee review of individual data. As a descriptive observational study, it was not registered; the study protocol is publicly available with the analysis code (see Data sharing). Reporting was guided by the STROBE statement and its RECORD extension^21,22^.

## Results

### National growth with prefectural variation in DBS management activity

National DBS management billings increased over 11 years, from 18,659 to 27,233 between 2014 and 2024 (Figure 1b), with a cumulative total of 258,408 billings. The number of patients with at least one DBS management billing also increased nationally in the most recent years, from 5,249 to 5,535 between 2022 and 2024. To characterise the current geographic status, we mapped the 2020–2024 mean DBS management billing rate for each prefecture, which varied widely (median, 153.5 per million; IQR, 79.0 to 273.2; Figure 1a).

**Fig. 1.**
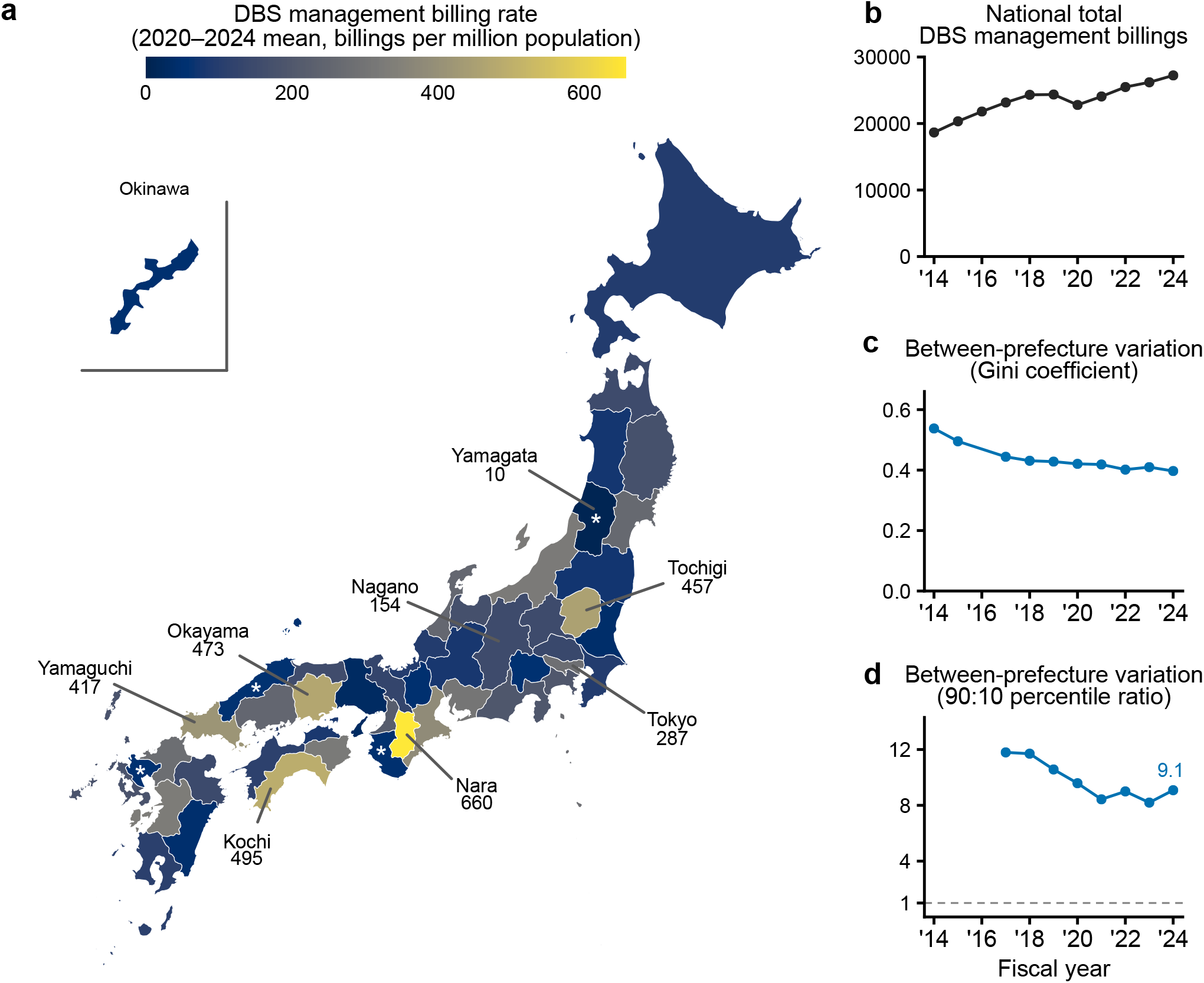
Recorded DBS management activity. **a)** Recent geographic distribution of DBS management billing rate (mean of available 2020–2024 values; asterisks in the map mark Yamagata, Saga, Shimane, and Wakayama, which are based on fewer than five years because of suppression, see Methods). **b)** National growth in DBS management billings. A transient decline in 2020 (*−*6.4% versus 2019) coincided with the COVID-19 pandemic. **c)** Annual between-prefecture variation in DBS management billing rates, by the Gini coefficient (0 indicates perfect equality; 1 indicates perfect inequality). **d)** The same variation by the 90:10 percentile ratio; the dotted line marks a ratio of one. c and d omit fiscal years for which the measure could not be estimated because of small-cell suppression, including 2016 (all prefectures suppressed, c and d) and 2014–2015 (seven suppressed prefectures, d).

Between-prefecture unevenness in billing rates remained substantial throughout the study period: in 2024, the prefecture at the 90th percentile of the billing rate recorded 9.1 times that of the prefecture at the 10th percentile (Figure 1d), and the Gini coefficient ranged from 0.40 to 0.54 over the 11 years (Figure 1c; 0 indicates perfect equality; 1 indicates perfect inequality). By comparison, the equivalent Gini co-efficient (2018–2024 mean) was considerably lower for the workforce of neurologists (0.18), neurosurgeons (0.11), and physicians overall (0.09), indicating that the DBS management billing rate is distributed more unevenly than the underlying physician workforce. The unevenness in DBS management billing rates narrowed over time, but the narrowing decelerated: the Gini coefficient’s annual decrease averaged 0.022 per year in 2014–2019 versus only 0.006 per year in 2019–2024 (Figure 1c, Supplementary Table 1).

The recent geographic picture and its unevenness were both reproduced using population denominators intended to better reflect clinical need, and using alternative unevenness measures (Supplementary Figures 1, 2, and 5a-c).

**Fig. 2.**
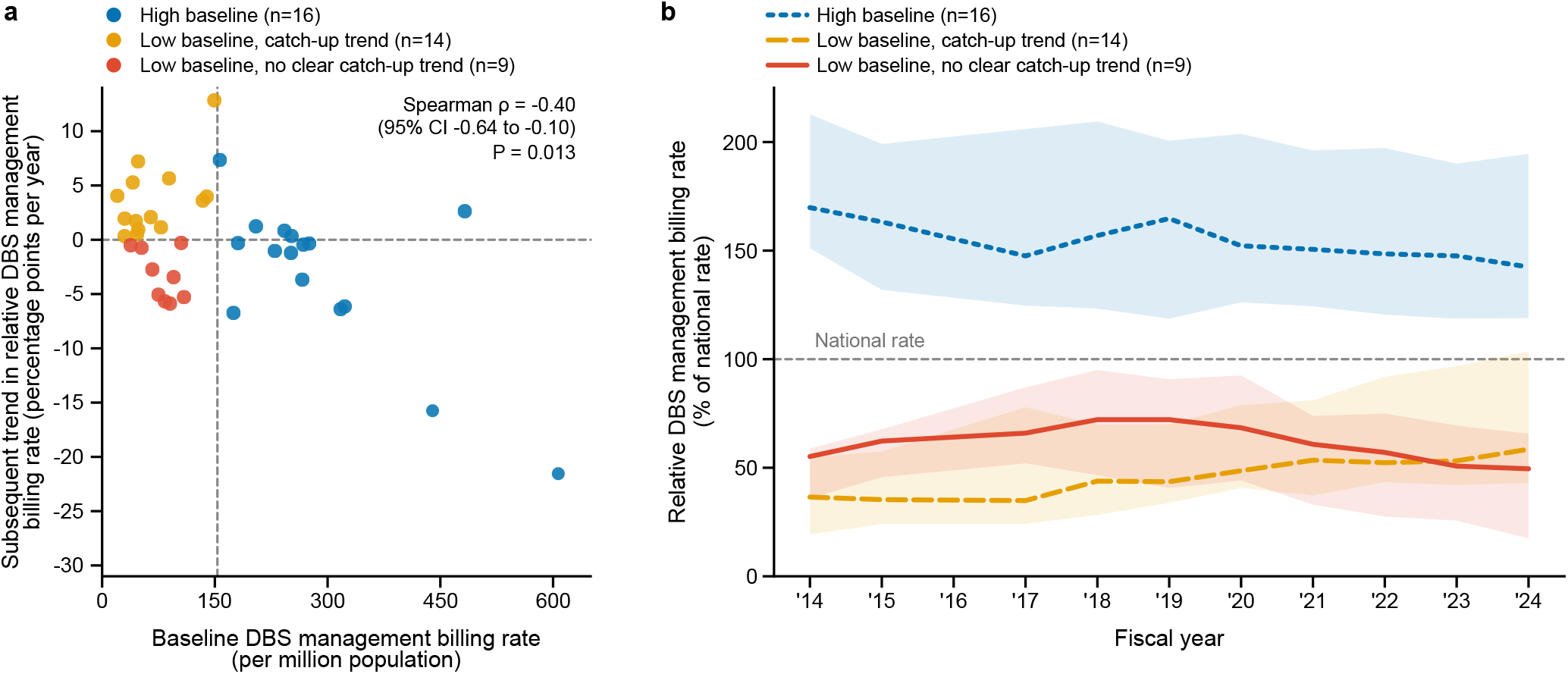
Longitudinal trend in relative DBS management billing rate. **a)** Baseline DBS management billing rate (2014–2015) and subsequent trend in relative DBS management billing rate (2017–2024). The analysis includes 39 prefectures with both baseline values and subsequent trend. The vertical dashed line marks the mean national baseline DBS management billing rate (153.4 per million population), and the horizontal dashed line marks a relative trend of zero. Together the two lines define the three groups used throughout panels a and b: blue, high baseline; orange, low baseline with a catch-up trend; red, low baseline with no clear catch-up trend. **b)** Median relative DBS management billing rate over 2014–2024 for the same three groups, with shading showing the interquartile range and the dashed line marking the national rate. Fiscal year 2016 is omitted because all prefecture-level billing counts were suppressed that year.

### Longitudinal trends in DBS management activity

The persistent unevenness in DBS management billing rates reflects differences accumulated over many years, not recent dynamics of change. We therefore calculated a robust longitudinal trend in each prefecture’s billing rate relative to the national rate from 2014 through 2024 (Supplementary Figure 3a–b). Prefecture-specific trends ranged from *−*21.6 to +10.1 percentage points of the national rate per year. The relative billing rates, and their trends, closely tracked the corresponding relative rates and trends of patients who received DBS management over 2022–2024 (Spearman *ρ* = 0.95 [95% CI 0.88 to 0.97] and 0.72 [95% CI 0.49 to 0.87], respectively; Supplementary Figure 4a–b).

**Fig. 3.**
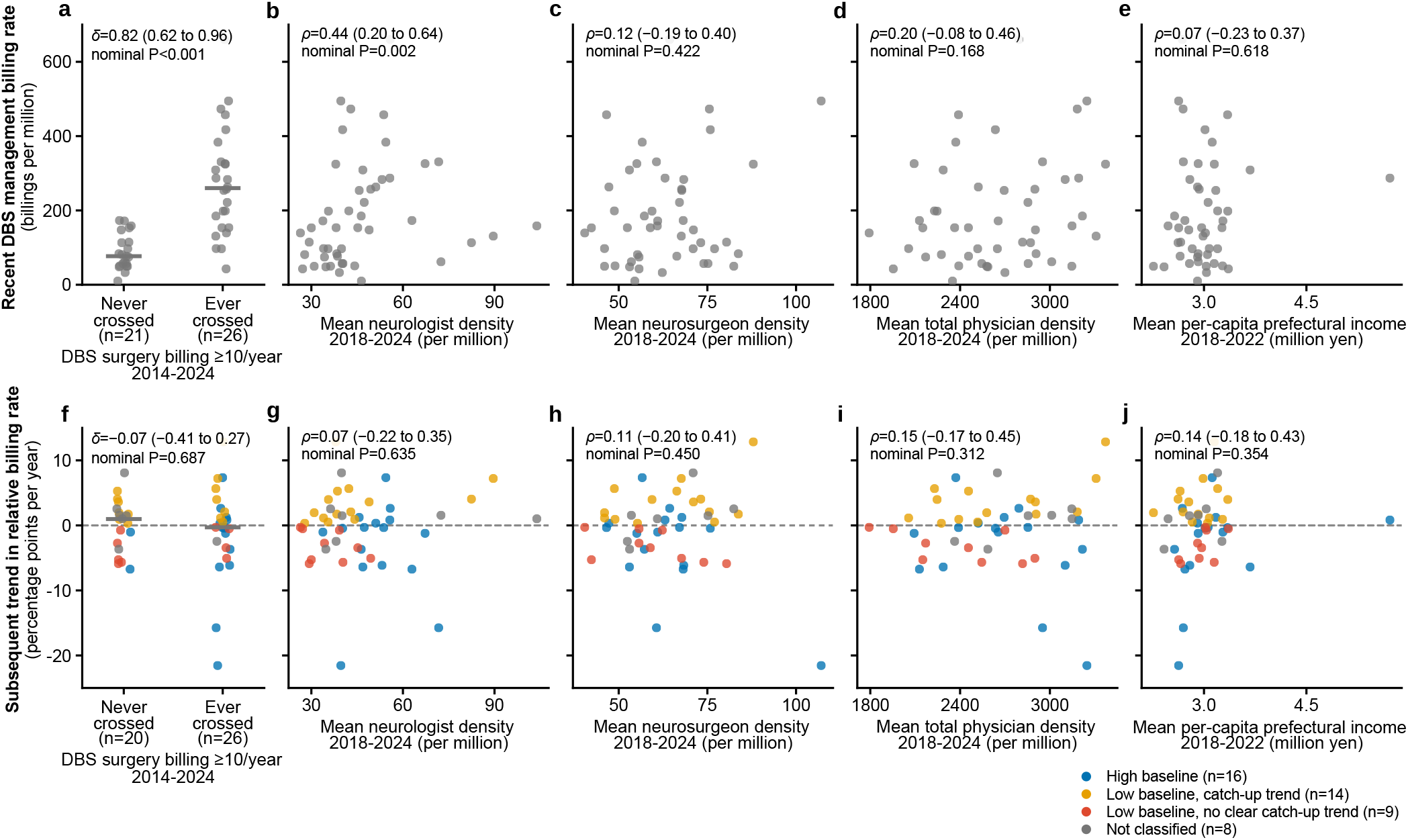
Clinical infrastructure and economic correlates of DBS management billing rate. **a-e)** Recent DBS management billing rate (mean 2020–2024) by a) DBS surgery billing (never versus at least once crossing the public reporting threshold, 2014-2022), b) mean neurologist density, c) mean neurosurgeon density, d) mean total physician density (per million population, 2018-2024 mean), and e) mean per-capita prefectural income (2018-2022 mean). **f-j)** The subsequent relative trend (2017-2024, as in Figure 2) against the same five indicators in the same order. Panels within each row (a-e, f-j) share the same y-axis scale. Dots show individual prefectures; point colours in f-j follow the Figure 2 trajectory groups; grey marks prefectures that could not be classified into a Figure 2 group (missing baseline or subsequent trend data). Bars in a and f show medians. *δ* is Cliff’s delta (panels a, f); *ρ* is Spearman’s rho (panels b-e, g-j); parenthetical values are 95% CI. P values from a two-sided permutation test (panels a, f) or from Spearman correlation with a two-sided permutation test (panels b-e, g-j).

Each prefecture’s trend reflects its own history of DBS management growth, enabling us to assess whether low-baseline prefectures are catching up towards the national average. Among the 23 complete-case prefectures with a baseline billing rate below the national mean, nine had a negative subsequent trend and therefore showed no clear catch-up trend (red points in Figure 2a, Supplementary Table 2). This categorisation was reproduced using a population denominator intended to better reflect clinical need (Supplementary Figure 5d, Supplementary Table 2). Across all 39 prefectures with complete baseline data, however, a lower baseline billing rate was overall associated with a more positive subsequent relative trend (*ρ* = *−*0.40; 95% CI, *−*0.64 to 0.10; P = *−*0.013; Figure 2a) (Supplementary Figure 5d, Supplementary Tables 3 and 4).

The corresponding trajectories of the relative DBS management billing rate are shown in Figure 2b. Prefectures showing no clear catch-up trend (red solid line) remained at approximately half the national rate throughout the period (median 50% [IQR 17–66%] of the national rate in 2024). The other two groups converged towards the national rate; nevertheless, the catch-up group (yellow dashed line) remained at only me-dian 59% (IQR 43–104%) of the national rate in 2024.

### Clinical infrastructure and economic correlates of DBS management activity

Finally, in an exploratory analysis, we related recent DBS management billing rate, and its subsequent relative trend, to clinical infrastructural and economic indicators (Figure 3). For the clinical infrastructure, recent billing rate was higher in prefectures with recorded DBS surgery billing (Cliff’s *δ* = 0.82; 95% CI, 0.62 to 0.96; Figure 3a) and increased with neurologist density (*ρ* = 0.44; 95% CI, 0.20 to 0.64; Figure 3b), but was not associated with neurosurgeon or total physician density (Figure 3c, d). None of the four indicators was associated with the subsequent relative trend (Figure 3f–i). Beyond clinical infrastructure, regional economic strength, represented by per-capita prefectural income, was not associated with either recent billing rate or its subsequent trend (Figure 3e, j).

## Discussion

### Principal findings

The long-term benefits of DBS treatment for chronic neurological disorders depend on ongoing management, and the geographic distribution of the management activity reflects the infrastructure available for sustained follow-up. Yet it has rarely been examined nationally. Our analysis of near-complete Japanese national claims showed that DBS management billings increased over 11 years, while prefectural billing rates remained markedly uneven and persistent. Although lower baseline billing rates were associated with more positive subsequent relative trends overall, this national-level association concealed that nine of the 23 low-baseline prefectures showed no clear catch-up trend through the period. To our knowledge, no comparable annual, region-level description of ongoing DBS management exists elsewhere.

### Geographic disparities in DBS treatment provision

Regional variation in DBS implantation surgery provision, in contrast, has been documented in several healthcare systems. A two-year census in Canada described provincial surgical rates ranging from 32% to 374% of the national average^23^; national analyses in the United States and China each estimated significant macro-regional differences in DBS surgical activity^10,24^. Our findings add a further stage to this sequence of DBS treatment provision: unevenness in ongoing management may compound the unevenness already documented in implantation, shaping the overall geographic pattern.

Geographic unevenness likely traces back to causal factors, though disentangling them is difficult beyond an exploratory level. A report from Canada linked neurosurgeon workforce availability to provincial rates on implantation surgery^23^, paralleling the neurologist-density association we found for on-going management, suggesting that specialised workforce availability underlies unevenness at both stages of DBS treatment provision. Economic strength has also repeatedly been linked to greater DBS surgical access, both at the individ-ual level in North America and at the macro-regional level in China^10,23,24^. We found no such association between region-level economic strength and DBS management measures. Whether ongoing management is as sensitive as surgery to economic strength remains unclear, but Japan’s universal healthcare system may itself help explain this difference, since it narrows disparities in the treatment options available to individuals.

### Uneven service delivery despite universal health coverage

Persistent unevenness of DBS management activity under Japan’s universal insurance and reimbursement framework illustrates the distinction between formal coverage and geographic service delivery. Similar prefectural unevenness has been reported for cardiac rehabilitation, radiotherapy, and home oxygen therapy under the same framework^19,25,26^, which, together with our findings, suggests that universal health coverage should be complemented by policies that ensure geographically equitable service delivery^27^. Moreover, the overall baseline–trend association concealed that nine low-baseline prefectures showed no clear catch-up trend, highlighting the need for prefecture-level evaluation of policies intended to alter geographic service delivery, as already flagged in the government’s report^28^.

Not every specialist service may need to be geographically even; concentration at a few centres is often appropriate for episodic, highly specialised procedures such as implantation, given the need for optimal workforce distribution. However, ongoing DBS management differs: it is long-term care for a chronic disease that must be sustained over years, while DBS is a standard and routinely considered treatment option. Ensuring its geographically equitable delivery therefore requires explicit strategies.

### Potential strategies to reduce geographic unevenness

One potential strategy is to decouple long-term DBS management from the centres that perform implantation surgery. This direction is consistent with recent proposals to shift Japan’s health system from hospital-centred episodic delivery towards longitudinal care closer to patients’ communities^29^. In our exploratory analysis, recent management billing rate was indeed higher in prefectures with higher implantation surgical billing, consistent with management remaining concentrated at these centres. A possible precedent for such decoupling is the 2012 establishment of C110-2 billing itself, which formalised DBS management as a billable activity distinct from surgery, and may have contributed to the faster narrowing of unevenness in the earlier half of the study period. Reimbursement for remote DBS management from 2026 could extend this decoupling further^30,31^. Our observations through 2014–2024 provide a pre-implementation benchmark for evaluating whether this policy reduces geographic unevenness.

A complementary strategy may be to expand DBS management expertise beyond implantation centres. As programming becomes increasingly complex and technically demanding^32^, a hub-and-spoke system combining continued support from expert centres with validated remote technologies could bring routine management closer to patients. In our exploratory analysis, recent management billing rate was associated with neurologist density, whereas subsequent regional growth was not associated with any measured physician-workforce indicator, highlighting the need to measure DBS-specific service capacity directly, including the availability of clinicians actively providing DBS management.

### Limitations

#### Several limitations warrant careful consideration

Some potential sources of bias were checked directly and did not change the conclusions. The primary measure counts billings rather than distinct patients, but prefecture-level relative billing rates closely tracked actual relative patient rates in available data (2022–2024). Diagnosis is unavailable, and the total-population denominator does not directly represent clinical need, but alternative denominators — adults aged 65 years and older, a certificate-based proxy for the advanced-PD population, and sex- and age-standardised claim ratios — gave similar results and the same conclusions (Supplementary Figures 1 and 5).

Other limitations would be expected to bias towards under-estimating the true extent and persistence of geographic unevenness. Small-cell suppression disproportionately removes the lowest tier of prefecture-years from the longitudinal analyses. The count of prefectures without a clear catch-up trend is conservative for two reasons: the catch-up assignment itself is permissive, since any positive subsequent trend counts as catch-up; and the descriptive baseline–trend association cannot rule out regression to the mean, which would bias low-baseline prefectures towards artefactually more positive subsequent trends.

Conversely, some limitations could lead us to overestimate patient burden if regional delivery patterns are used to infer individual access. Prefecture identifies the location of the billing institution rather than the patient’s residence, so low local activity could reflect efficient referral to a nearby centre rather than an individual access gap. Nevertheless, unlike the episodic implantation procedure, ongoing management requires repeated contact over many years. Geographic concentration of its delivery may therefore impose a meaningful travel burden, consistent with interpretations of prefecture-level unevenness in other recurring services in Japan^25,26^.

Finally, other limitations have no predictable direction. DBS is known to be underused among potentially eligible patients^5^, but our claims analysis captures only delivered care and therefore cannot identify unmet need. This study is descriptive and could not infer why this unevenness exists; the exploratory infrastructural associations rely on metrics available from open data sources and are therefore not comprehensive. The ongoing expansion of remote management may offer an opportunity for a natural experiment that could partially clarify its causes.

Taken together, the observed geographic unevenness and its persistence were robust, although its implications for individual access remain unconfirmed. We therefore state our conclusions at the level of the health system, where the data inform directly, and treat implications for individual access as a hypothesis for future patient-level studies.

### Conclusion

In conclusion, recorded ongoing DBS management activity was geographically uneven across Japan. Our observations suggest that universal health coverage and national-level growth do not guarantee equitable geographic delivery of long-term DBS management. These nationwide, longitudinal observations can inform policies aimed at reducing geographic barriers to long-term specialist care across healthcare systems.

## Supplemental Information

The Supplementary Materials (Supplementary Figures 1–5, Supplementary Tables 1–4, the Supplementary Methods, and the Supplementary Code Guide) are appended.

## FUNDING

No specific funding was received for this work.

## ACKNOWLEDGEMENTS

We thank the volunteers and authors of Natural Earth for providing free and open public-domain map data, and the developers of the Okabe-Ito colour-blind-friendly colour palette for developing and publicly sharing it for use throughout our figures.

## COMPETING INTERESTS

The authors declare no competing interests.

## DATA AND CODE AVAILABILITY

All input data are publicly available from the sources listed in the study protocol and source manifest. The source manifest, processed analytic data, complete analysis code, and study protocol are available in Harvard Dataverse (DOI: 10.7910/DVN/FJNBCK).

## GENERATIVE AI USAGE

The authors used ChatGPT (OpenAI) and Claude (Anthropic) to assist with language editing, and Cursor (Anysphere), an AI-assisted code editor with automatic model selection enabled, to assist with analysis code development. The authors reviewed and revised all resulting content and take full responsibility for the final manuscript.

## AUTHOR CONTRIBUTIONS

JT and TH conceived and designed the study. JT collected and curated the data, developed and performed the analyses, and produced the figures. TH reviewed the source data and analytical outputs and supervised the study. Both authors interpreted the findings and directly accessed and verified the underlying data. JT drafted the manuscript, and TH critically revised it. Both authors confirm that they had full access to all the data in the study, approved the final version, and accept responsibility to submit for publication.

## Supplementary Materials

**Fig. S1.**
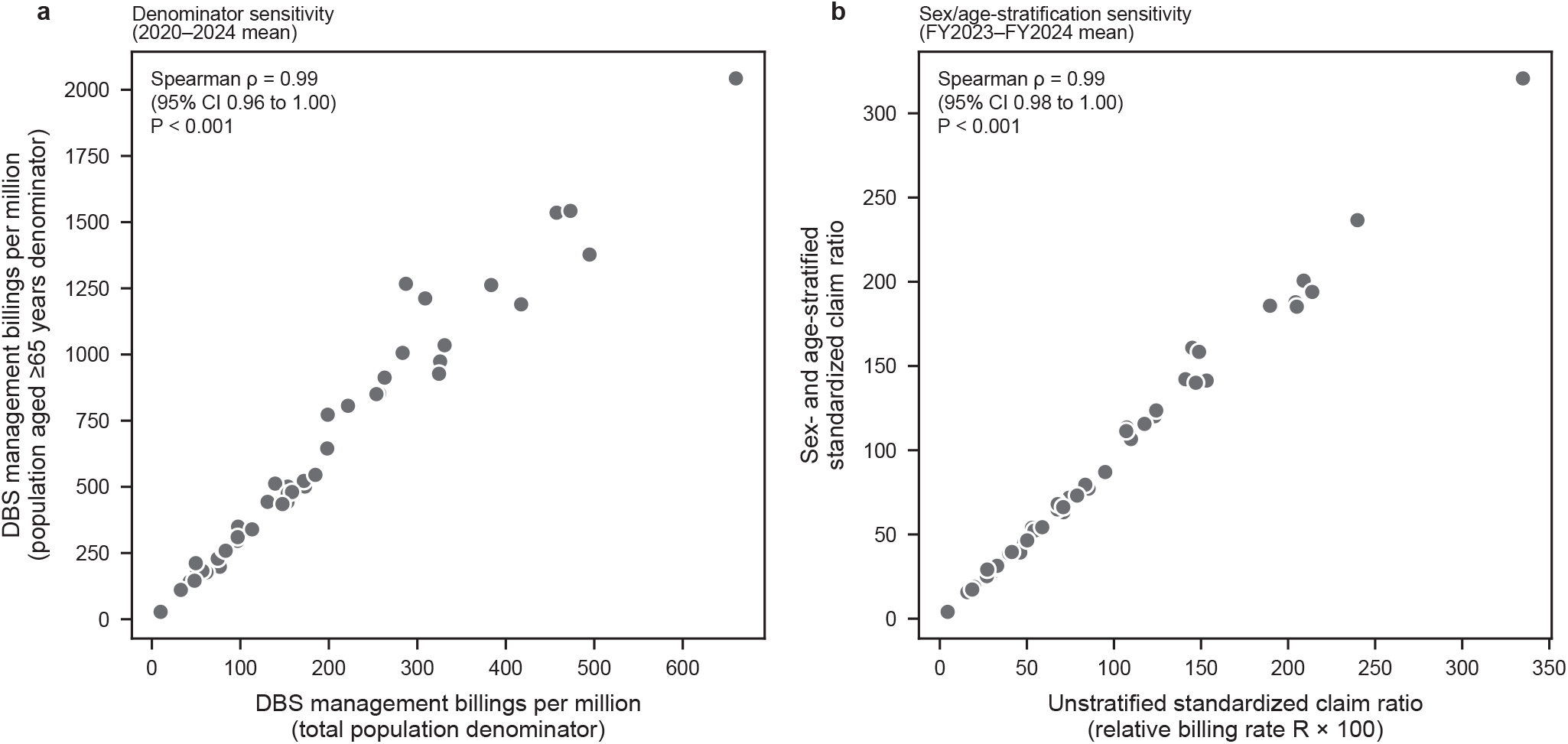
DBS management billing rate under alternative denominators and standardization. **a)** 2020–2024 mean billing rate for each prefecture (n = 47), using the total population versus the population aged *≥*65 years as denominators (year-matched before averaging). **b)** Prefecture standardized claim ratio (SCR; see Methods), unstratified versus stratified by sex and age, for 2023–2024 (mean of available years). Spearman correlation shown in each panel. 95% CIs were based on 20,000 prefecture-level bootstrap resamples; P values were based on a two-sided permutation test with 100,000 permutations.

**Fig. S2.**
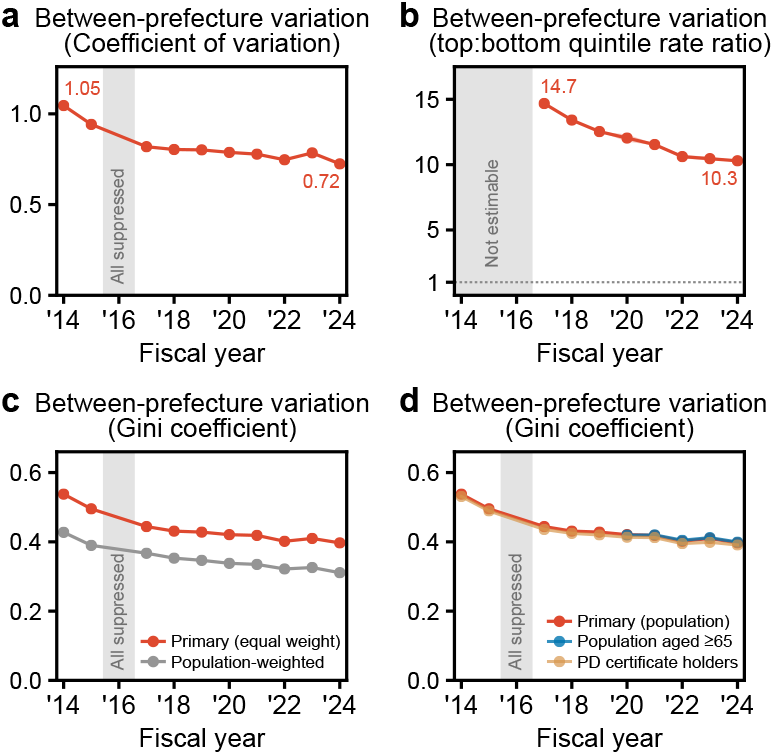
Alternative measures of between-prefecture variation. Annual between-prefecture variation in billing rates, by a) Coefficient of variation, and b) Ratio of the summed billing rate in the nine highest-rate prefectures to that in the nine lowest; in b, the dotted line marks a ratio of one. c, d) The primary Gini coefficient of Figure 1c was compared with c) a population-weighted rather than equal prefecture weights (primary), and d) variants using population aged *≥*65 years or PD certificate holders rather than the total population (primary) as the denominator (the population aged *≥*65 years is available only from 2020). Shading shows the range consistent with the suppressed cells, and grey bands mark fiscal years in which the measure cannot be estimated (see Supplementary Methods).

**Fig. S3.**
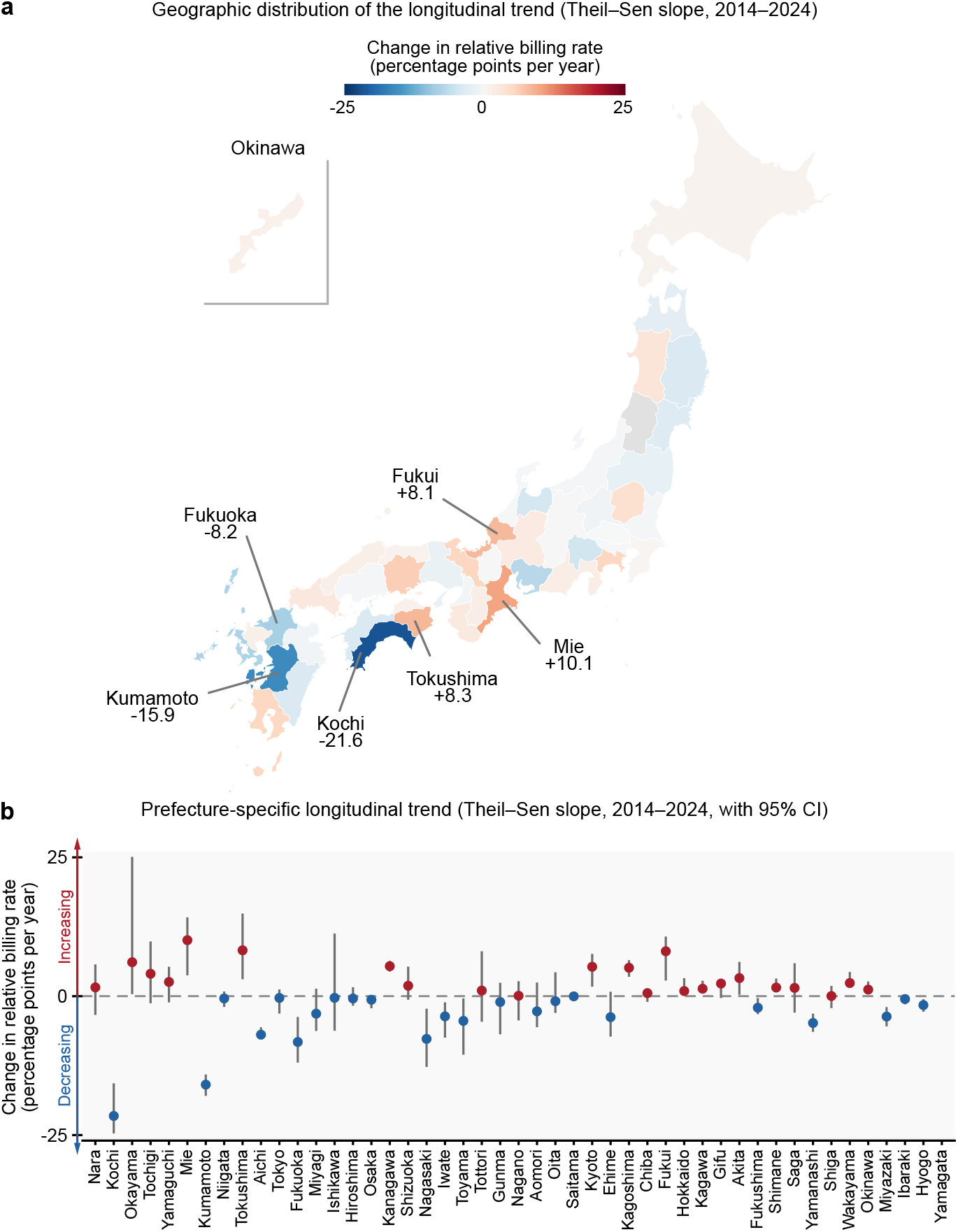
Prefecture-specific longitudinal trend in relative DBS management billing rate. **a)** The 2014–2024 prefecture-specific Theil–Sen trends are mapped (red = positive trend, blue = negative trend). **b)** The same trends by prefecture, with rank-based 95% confidence intervals. In a and b, trends are shown only for prefectures with at least six available annual values.

**Fig. S4.**
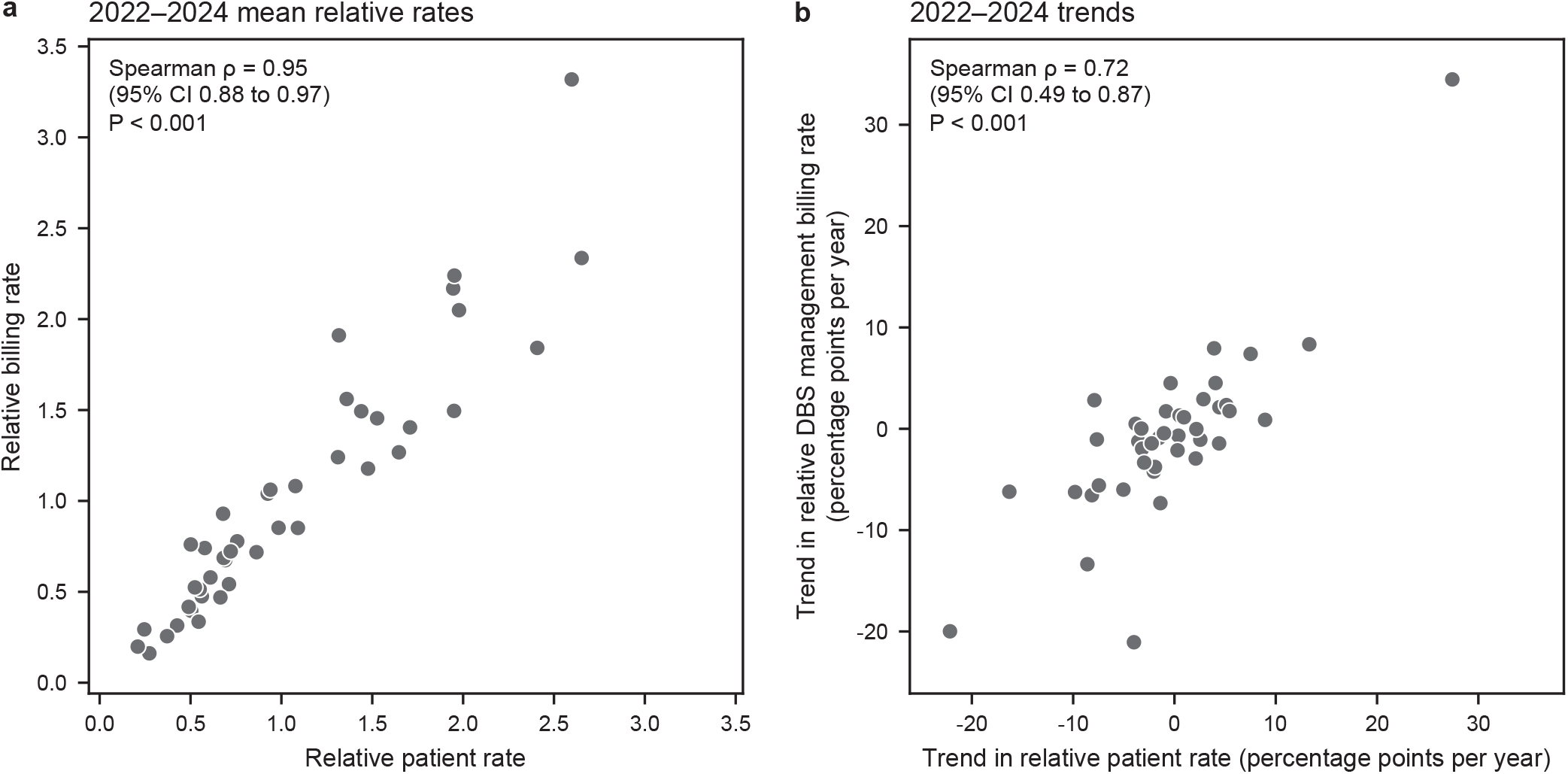
Validation of DBS management metrics against managed-patient counts. The patient rate (ID5-linked, see Methods): the annual number of patients receiving DBS management per million population. **a)** 2022–2024 mean relative patient rate compared with the corresponding mean relative DBS management billing rate among prefectures with complete three-year data (n = 42). **b)** The FY2022–FY2024 Theil–Sen trend in the relative patient rate compared with the corresponding trend in the relative DBS management billing rate among prefectures with complete three-year data (n = 42). Relative rates are prefecture rates divided by the corresponding national rate in the same fiscal year. Nationally, C110-2 billings per ID5-linked patient were stable across 2022–2024 (4.85–4.92). 95% CIs were based on 20,000 prefecture-level bootstrap resamples; P values were based on a two-sided permutation test with 100,000 permutations.

**Fig. S5.**
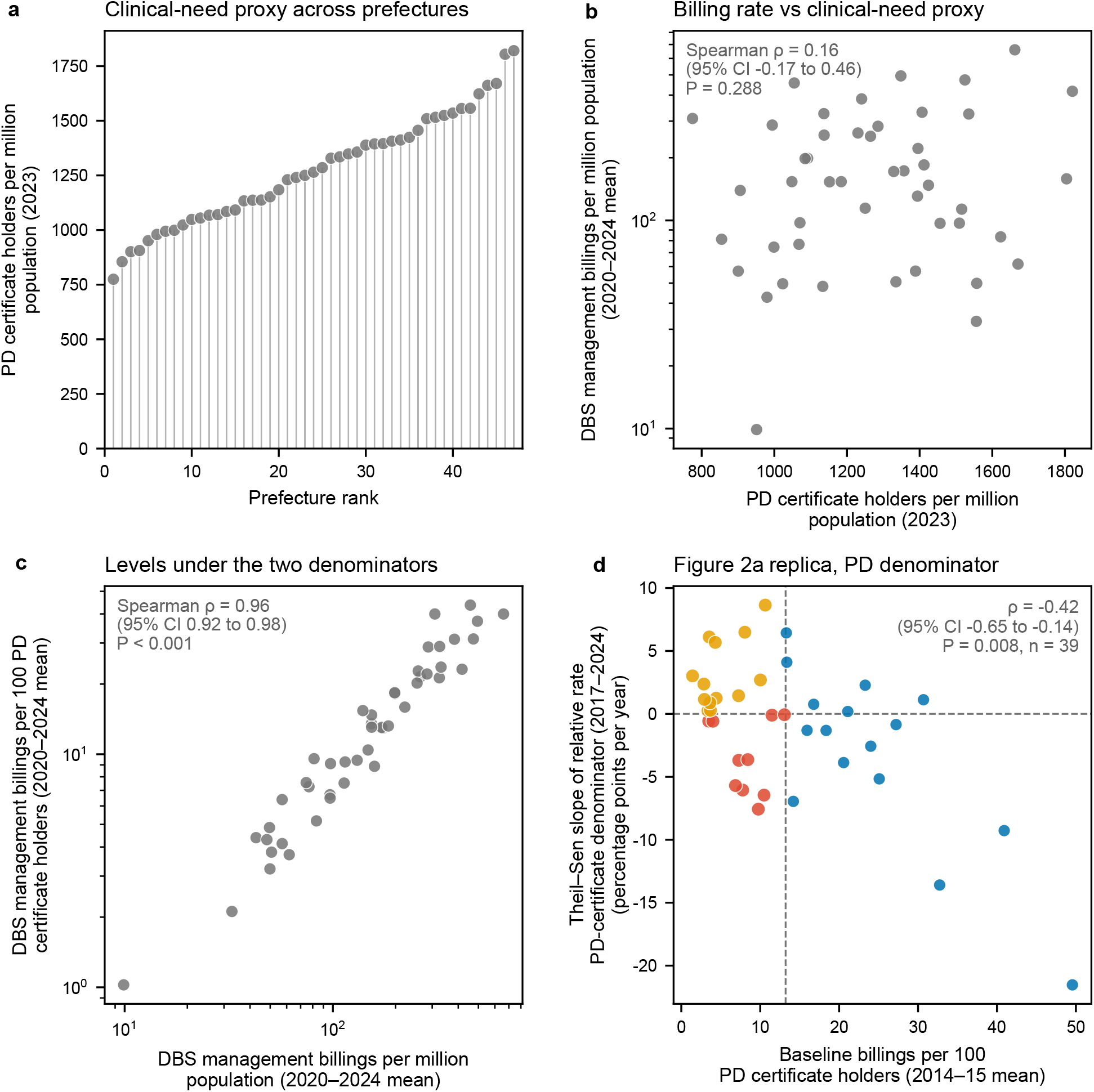
DBS management billing rates using the total population versus a Parkinson’s disease certificate-based denominator. Robustness of the main findings to replacing the population denominator with an approximation of the advanced-PD population: holders of a specified medical care certificate for Parkinson’s disease (see Supplementary Methods). **a)** Certificate holders per million population across the 47 prefectures, ranked from lowest to highest. **b)** 2020–2024 mean DBS management billing rate against the certificate rate, indicating that the geographic pattern of management billing rates is not shaped by the distribution of PD certificate holders. **c)** Direct comparison of the 2020–2024 mean billing rate (per million population) against the 2020–2024 mean billing rate (per 100 PD certificate holders): the two denominators gave a closely rank-correlated ordering of prefectures. **d)** Replica of the Figure 2a analysis using certificate denominators: the baseline is the 2014–2015 mean number of billings per 100 certificate holders and the outcome is the 2017–2024 trend of the prefecture-to-national ratio of billings per certificate holder. Point colours follow Figure 2a. The vertical dashed line marks the national baseline (13.2 billings per 100 certificate holders). Twenty-two of the 23 low-baseline prefectures in the primary analysis remained low-baseline under the certificate denominator, and the catch-up categorization itself was unchanged for all 23 prefectures regardless of denominator. 95% CIs were based on 20,000 prefecture-level bootstrap resamples; P values were based on a two-sided permutation test with 100,000 permutations.

**Supplementary Table 1.** Sensitivity of the Gini-coefficient deceleration estimate to the choice of split year. The Results report that the narrowing of the Gini coefficient (Figure 1c) decelerated, based on the average annual decrease before versus after 2019. This table repeats that calculation for every split year from 2017 to 2022, each row using the same full-study-period endpoints (2014, 2024) and the same Gini values underlying Figure 1c. The direction of the finding — a faster decrease before the split than after it — did not depend on the

| Split year | Period before split (FY) | Annual Gini decrease<br>before split (per year) | Period after split (FY) | Annual Gini decrease<br>after split (per year) |
| --- | --- | --- | --- | --- |
| 2017 | 2014–2017 | 0.031 | 2017–2024 | 0.007 |
| 2018 | 2014–2018 | 0.027 | 2018–2024 | 0.006 |
| 2019* | 2014–2019 | 0.022 | 2019–2024 | 0.006 |
| 2020 | 2014–2020 | 0.020 | 2020–2024 | 0.006 |
| 2021 | 2014–2021 | 0.017 | 2021–2024 | 0.007 |
| 2022 | 2014–2022 | 0.017 | 2022–2024 | 0.002 |

\*Split year used in the main text.

**Supplementary Table 2.**
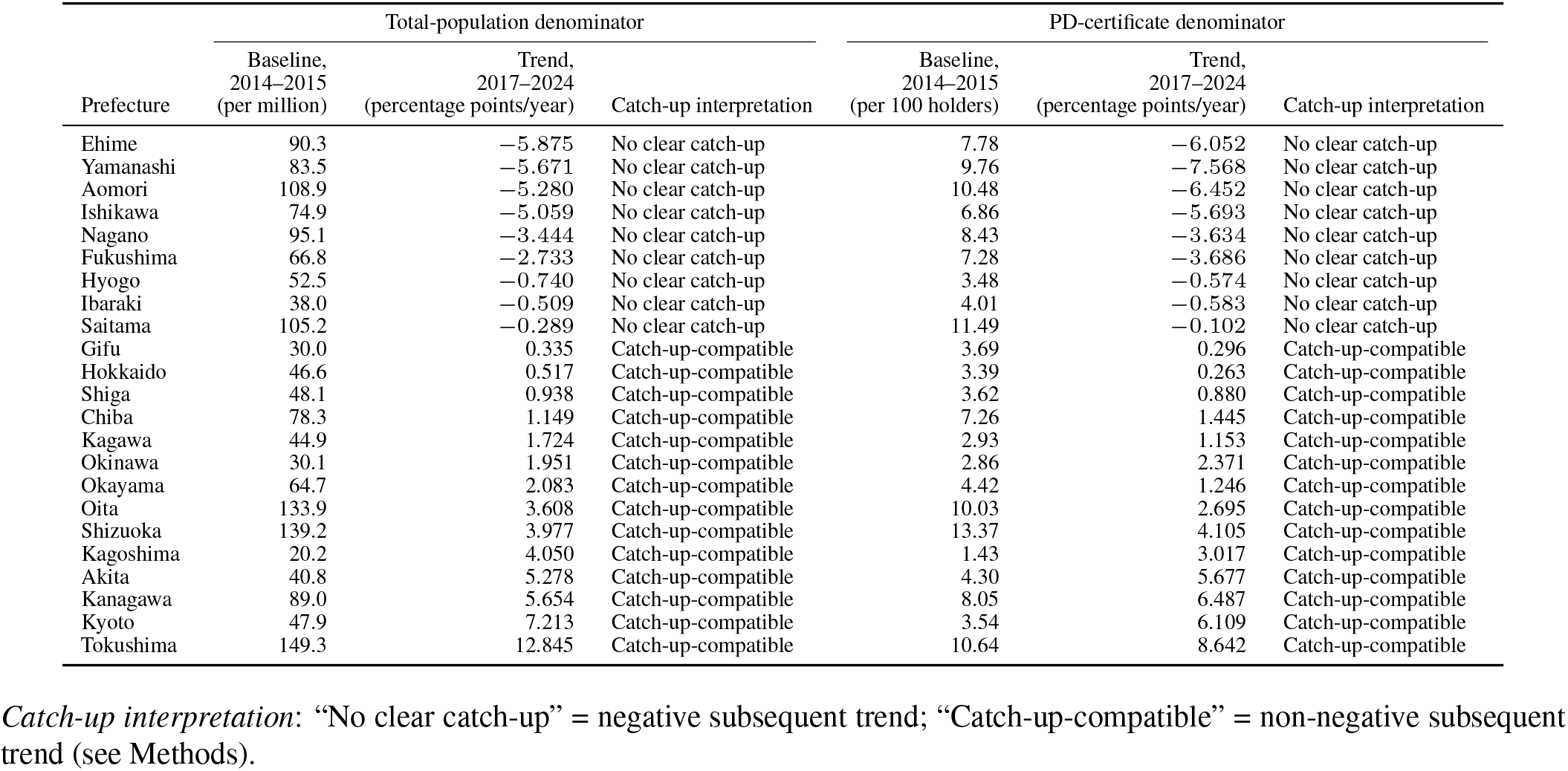
Low-baseline prefectures and subsequent relative trends, under the total population and PD-certificate denominators. The 23 complete-case prefectures with a 2014–2015 mean billing rate below the corresponding national mean of 153.4 billings per million population. Catch-up classification follows the definition in Methods (a negative subsequent trend indicates no clear catch-up trend). The three rightmost columns repeat this classification using the Parkinson’s disease certificate denominator described in Supplementary Figure 5, for the same 23-row set. Two prefectures’ low-baseline status is not concordant between denominators (Supplementary Figure 5): Osaka falls below the national baseline only under the certificate denominator and so is not part of this table; Shizuoka, included here, has a certificate-denominator baseline (13.4 per 100 certificate holders) just above the certificate-denominator national baseline (13.2) and so is not classified as low-baseline under that denominator.

**Supplementary Table 3.** Overall baseline–trend association under alternative baseline definitions. Comparison with the complete-case 2014–2015 mean baseline used in Figure 2a, under three alternative definitions: 2014 alone, 2015 alone, and the mean of whichever baseline-year values were available (see Supplementary Methods). Each row reports the number of prefectures, Spearman *ρ*, 95% prefecture-bootstrap confidence interval, and two-sided permutation P value.

| Analysis | Baseline definition | *n* | Spearman $\rho$ [95% CI] | *P* |
| --- | --- | --- | --- | --- |
| Primary | 2014–2015 complete-case mean | 39 | −0.40 [−0.64 to −0.10] | 0.013 |
| Sensitivity | 2014 only | 40 | −0.38 [−0.64 to −0.08] | 0.014 |
| Sensitivity | 2015 only | 40 | −0.39 [−0.64 to −0.09] | 0.014 |
| Sensitivity | Mean of available baseline years | 41 | −0.40 [−0.64 to −0.10] | 0.010 |

**Supplementary Table 4.** Overall baseline–trend association with the trend period truncated at 2022. Comparison with the 2017–2024 trend used in Figure 2a, ending instead at 2022, before DBS reimbursement was extended to drug-resistant focal epilepsy in December 2023 (see Supplementary Methods). Both analyses use the complete-case 2014– 2015 mean billing rate as the baseline. Each row reports the number of prefectures, Spearman *ρ*, 95% prefecture-bootstrap confidence interval, and two-sided permutation P value.

| Analysis | Trend period | *n* | Spearman $\rho$ [95% CI] | *P* |
| --- | --- | --- | --- | --- |
| Primary | 2017–2024 | 39 | −0.40 [−0.64 to −0.10] | 0.013 |
| Sensitivity | 2017–2022 | 39 | −0.34 [−0.63 to 0.00] | 0.034 |

## Supplementary Methods: Statistical Analysis

### Alternative population denominators for the DBS management billing rate

As the primary analysis, the DBS management billing rate was calculated using the total population per prefecture as the denominator (Statistics Bureau of Japan population estimates [M1]; see Methods). Robustness to this population denominator was assessed using two alternative denominators, recalculated for each available FY2020–FY2024 prefecture-year: the population aged 65 years or older, and the number of holders of a specified medical care certificate for Parkinson’s disease (PD) at the end of FY2023 (Health Administration Reports). We used this certified-PD denominator because PD is the predominant DBS indication and certification requires Hoehn and Yahr stage 3 or higher with impaired activities of daily living, so certificate holders approximate the advanced-PD population driving most DBS management demand. A single fixed reference year (FY2023) was used for the PD-certificate denominator because prefecture-level counts were unavailable for FY2016, and counts before FY2015 combined Parkinson’s disease with other parkinsonian syndromes under the former disease category. These denominators were used in Supplementary Figure 1a (population aged *≥*65 years), Supplementary Figure 5 (PD-certificate denominator), Supplementary Figure 2d (Gini coefficient recomputed under both alternative denominators), and Supplementary Table 2 (low-baseline classification under the PD-certificate denominator).

### Alternative measures of between-prefecture variation in the DBS management billing rate

As the primary analysis, between-prefecture unevenness in the billing rate was summarized by the Gini coefficient and by the ratio of its 90th to its 10th percentile (Figure 1c–d; see Methods). It was also summarised by the coefficient of variation and by the ratio of the summed billing rate in the nine highest-rate prefectures to that in the nine lowest (the top and bottom fifths) (Supplementary Figure 2a–b). Each measure was calculated over all 47 prefectures by back-calculating suppressed cells from their recoverable total, reported as the resulting interval; this interval is shaded in Supplementary Figure 2, while Figure 1c–d shows only the central estimate and omits fiscal years that could not be estimated (see Figure 1 legend). FY2016 could not be estimated for any measure because all prefecture values were suppressed that year; in addition, the 90:10 percentile ratio could not be estimated for FY2014–FY2015, when seven simultaneously suppressed prefectures left its bounds too wide to be informative. The Gini coefficient was also recomputed weighting prefectures by population, and using the population aged 65 years or older and the number of PD certificate holders as denominators (Supplementary Figure 2c–d).

### Sex- and age-stratified standardized claim ratio versus the unstratified rate

As the primary analysis, the DBS management billing rate was expressed relative to the national rate, equivalent (*×*100) to a conventional unstratified standardized claim ratio (SCR; see Methods). We also assessed the robustness of this metric to sex- and age-stratification (Supplementary Figure 1b). National counts by sex and five-year age band were taken from the NDB Open Data sex-and-age tables, and prefecture populations in the corresponding strata (sex *×*age 0–14, 15–64, 65–74, and 75 years or older) from the Statistics Bureau population estimates [M1]. These data are available only for FY2023 and FY2024; each prefecture’s value is the mean of the available values across the years.

### Overall baseline–trend association under alternative baseline definitions and trend period

As the primary analysis, each prefecture’s FY2014–FY2015 mean baseline billing rate was related to its subsequent FY2017– FY2024 trend (Figure 2a; see Methods). To assess the effect of this complete-case baseline definition, the FY2014–FY2015 mean was recomputed using FY2014 alone, FY2015 alone, and the mean of whichever baseline-year values were available (Supplementary Table 3). The association was also assessed after truncating the trend period at FY2022, before DBS reimbursement was extended to drug-resistant focal epilepsy in December 2023 (Supplementary Table 4).

### Data sources for population and physician workforce statistics

Prefecture and national population denominators were taken from the Statistics Bureau of Japan’s annual population estimates for each October 1 [M1]. FY2015 and FY2020 prefecture populations, which coincide with Japan’s quinquennial Population Census, were taken from the corresponding Census totals rather than the intercensal estimate. Neurologist, neurosurgeon, and total physician counts by prefecture were obtained from the FY2018, FY2020, FY2022, and FY2024 rounds of the Physician, Dentist and Pharmacist Statistics, a biennial national survey conducted by the Ministry of Health, Labour and Welfare and published via the e-Stat portal [M2], and divided by the corresponding population estimate [M1] to express density per million population.

## Supplementary Code Guide: the Japanese billing codes used in this study

This guide describes the basis for selecting C110-2 and the definitions and suppression rule for the codes relevant to this study.

### Background

Japan has a universal health insurance system in which medical services are assigned nationally standardized billing codes [C1]. Since fiscal year (FY) 2014, the Ministry of Health, Labour and Welfare (MHLW) has released aggregated prefecture-level and national counts for individual codes through the National Database of Health Insurance Claims and Specific Health Checkups (NDB) Open Data [C2–C4]. These data provide broad national capture of insured DBS-related care, excluding claims billed solely under public funding.

### Basis for C110-2 as the primary DBS indicator

At its introduction in 2012, MHLW described the C110-2 fee as covering the checking and programming of an implanted brain stimulator, including adjustment of stimulation contacts, amplitude, pulse width, and frequency for DBS [C5]. The Pharmaceuticals and Medical Devices Agency (PMDA) currently explicitly defines the corresponding device as stimulating a specific deep-brain region [C6]. MHLW has also used prefecture-level C110-2 counts as an indicator of regional DBS management activity, and the FY2026 revision directly linked C110-2 to remote DBS programming [C7,C8]. These sources support C110-2 as the most appropriate publicly available claims indicator of ongoing DBS management.

C110-2 is a billing measure, without any link to patient or provider information: the underlying diagnosis, the provider’s specialty, or whether care was delivered within a specialized care system are not specified [C9]. Since December 2023 it may also capture DBS for drug-resistant focal epilepsy, so the final study years are not strictly movement-disorder-specific [C10]. We therefore interpret C110-2 as a claims measure of ongoing DBS management, predominantly for movement disorders. We assessed the robustness of our findings to this epilepsy expansion by truncating the trend period before FY2023 (Supplementary Table 4).

### Billing codes relevant to this study

#### Distinctions between C110-2 and related codes

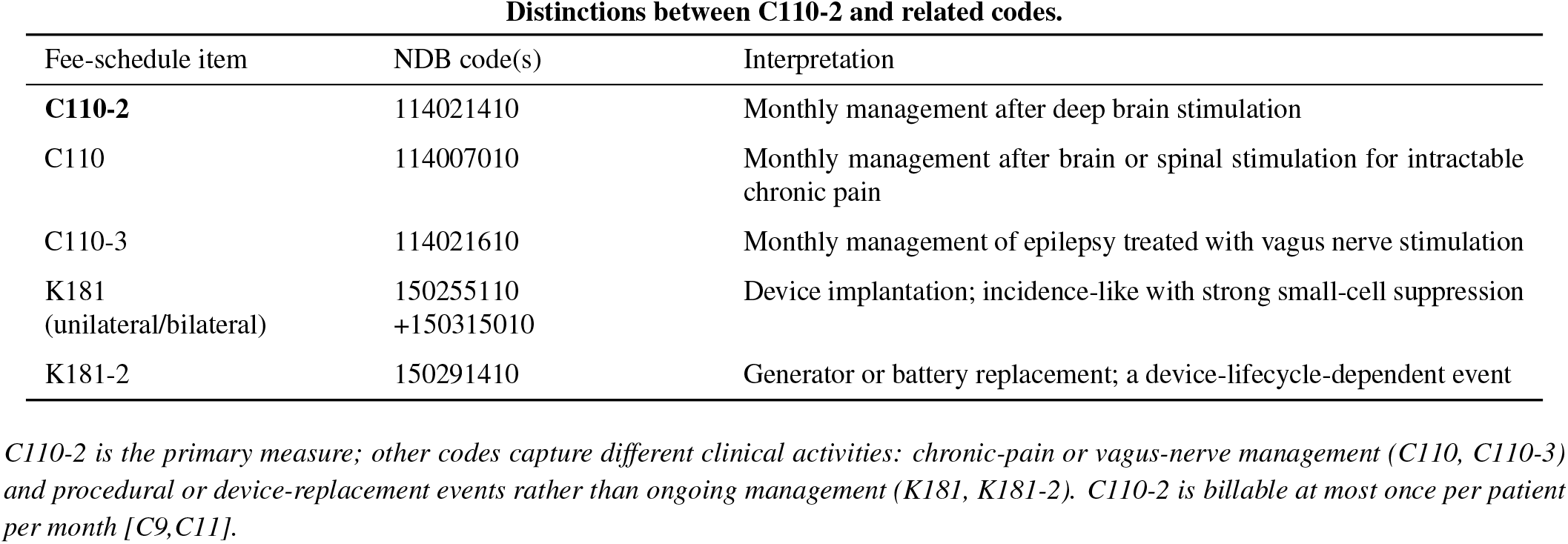

#### Small-cell suppression

A blank cell reflects a suppressed small count, not a true zero. Prefecture-year counts below 10 are suppressed and published as blank cells [C3]. When exactly one cell in a given year is suppressed, the entire set of prefecture values for that year is suppressed and only the total is shown. This rule accounts for the FY2016 all-prefecture suppression of C110-2 [C12,C13].

#### Patient counts for C110-2 claims (ID5)

For FY2022–FY2024 only, annual counts of patients with at least one C110-2 billing are available through ID5, a pseudonymous NDB identifier linking claims to the same patient. Prefecture-level patient counts follow the same suppression rule as billing counts. In this study, ID5-based counts were used solely to assess whether relative C110-2 billing rates tracked relative patient rates (Supplementary Figure 4).

#### K181 as the exploratory DBS-surgery indicator

The exploratory analysis (Methods; Figure 3 of the main text) used K181 (DBS device implantation; see the billing-code table above) as a binary indicator of whether a prefecture ever crossed the public reporting threshold for K181 billings during FY2014–FY2024, not as a quantitative measure of surgical volume.

## Notes

### Competing Interest Statement

The authors have declared no competing interest.

### Clinical Protocols

https://doi.org/10.7910/DVN/FJNBCK

### Author Declarations

All datasets used in this study were openly available before the reported analyses were initiated. We used only fully aggregated national- and prefecture-level data; no individual-level or identifiable information was accessed. The original data were obtained from the following publicly accessible sources: 1. Japanese Ministry of Health, Labour and Welfare, NDB Open Data, fiscal years 2014-2024: https://www.mhlw.go.jp/stf/seisakunitsuite/bunya/0000139390.html https://www.mhlw.go.jp/stf/seisakunitsuite/bunya/0000177221.html https://www.mhlw.go.jp/stf/seisakunitsuite/bunya/0000177221_00002.html https://www.mhlw.go.jp/stf/seisakunitsuite/bunya/0000177221_00003.html https://www.mhlw.go.jp/stf/seisakunitsuite/bunya/0000177221_00008.html https://www.mhlw.go.jp/stf/seisakunitsuite/bunya/0000177221_00010.html https://www.mhlw.go.jp/stf/seisakunitsuite/bunya/0000177221_00011.html https://www.mhlw.go.jp/stf/seisakunitsuite/bunya/0000177221_00012.html https://www.mhlw.go.jp/stf/seisakunitsuite/bunya/0000177221_00014.html https://www.mhlw.go.jp/stf/seisakunitsuite/bunya/0000177221_00016.html https://www.mhlw.go.jp/stf/seisakunitsuite/bunya/0000177221_00017.html 2. Population estimates and census data, Statistics Bureau of Japan and e-Stat: https://www.stat.go.jp/data/jinsui/ https://www.stat.go.jp/data/kokusei/ https://www.e-stat.go.jp/stat-search/file-download?fileKind=0&statInfId=000032142406 3. Specified-disease certificate counts and Physician, Dentist and Pharmacist Statistics, e-Stat: https://www.e-stat.go.jp/stat-search/file-download?statInfId=000040217273&fileKind=1 https://www.e-stat.go.jp/stat-search/files?toukei=00450026 4. Prefectural income, Cabinet Office of Japan: https://www.esri.cao.go.jp/jp/sna/data/data_list/kenmin/files/contents/tables/2022/soukatu7.xlsx 5. Geographic boundary data, Natural Earth: https://naturalearth.s3.amazonaws.com/10m_cultural/ne_10m_admin_1_states_provinces.zip A complete file-level source manifest, including the exact source URL for each of the 49 source files, is included in the replication package: https://doi.org/10.7910/DVN/FJNBCK

